# A CDE-based data structure for radiotherapeutic decision-making in breast cancer

**DOI:** 10.1101/2025.02.04.25321635

**Authors:** Fabio Dennstädt, Maximilian Schmalfuss, Johannes Zink, Janna Hastings, Roberto Gaio, Max Schmerder, Paul Martin Putora, Nikola Cihoric

**Author notes:** **Corresponding author:** Fabio Dennstädt, **mailing address:** Department of Radiation Oncology, Inselspital, Freiburgstrasse 18, 3010 Bern, SWITZERLAND.

## Abstract

**Background:** The increasing complexity and data-driven nature of oncology and radiation therapy necessitates structured and precise data management strategies. The National Institutes of Health (NIH) has introduced Common Data Elements (CDEs) as a uniform approach to facilitate consistent data collection. However, there is currently a lack of a comprehensive set of CDEs for describing situations for and within radiation oncology. Aim of this study was to create a CDE-based data structure for radiotherapeutic decision-making in breast cancer to promote structured data collection on the level of a local hospital.

**Methods:** Local Standard Operating Procedures (SOPs) were analyzed to identify relevant decision-making criteria used in clinical practice. Corresponding CDEs were identified and a structured data framework based on these CDEs was created. The framework was translated into machine-readable JSON format. Six clinical practice guidelines of the American Society for Radiation Oncology (ASTRO) were analyzed as full-text to investigate how many guideline recommendations and corresponding decision-making criteria could be presented using the data structure.

**Results:** The study identified 31 decision-making criteria mentioned in the SOPs, leading to the establishment of 46 CDEs. A hierarchical structure within an object-oriented data framework was created and converted into JSON format. 94 recommendations with mentioning of decision-making criteria in 216 cases were identified across the six ASTRO guidelines. In 151 cases (70.0%) the mentioned criterion could be presented with the data framework.

**Conclusions:** The CDE-based data structure represents a clear framework for structuring medical data for radiotherapeutic decision-making in breast cancer patients. The approach facilitates detailed description of individual breast cancer cases and aids in the integration of information technology. Furthermore, it promotes sharing of standardized data among healthcare providers.

## INTRODUCTION

The application of modern information technology (IT) systems and artificial intelligence (AI) is of ever increasing relevance in modern oncology and radiation oncology. For management of medical data within databases and for data processing within most IT applications, it is necessary to store the data in a structured and precise way. The concept of common data elements (CDEs), introduced in 2011 by the National Institutes of Health (NIH) for consistent data collection in clinical trials, can be used for this purpose as it facilitates clear definitions of semantic information. As explained in the definition, “a common data element is a standardized, precisely defined question, paired with a set of allowable responses, used systematically across different sites, studies, or clinical trials to ensure consistent data collection” (1), (2).

While the concept of using CDEs is already further advanced in medical disciplines like radiology (3) or neurology (4), currently there is a lack of a comprehensive list of defined data elements in radiation oncology (5).

Nevertheless, structured documentation using such a concept would be of high value in a highly technical discipline like radiation oncology – not only for research, but also for management of real-world data (RWD). The International Society for Radiation Oncology Informatics (ISROI) is therefore actively working on promoting the usage of CDEs in radiation oncology (6).

For clinical decision-making international guidelines as well as locally defined standard operating procedures (SOPs) are fundamental. While guidelines, SOPs and other sorts of literature regarding clinical decision-making are mentioning some sort of decision-making criteria, those are usually not presented in a formalized and clearly defined way (7). The clinical decision-making in breast cancer patients is particularly complex, as multiple factors and criteria are relevant while decision-making becomes more personalized and tailored to individual circumstances. As a result, a variety of IT-based tools and clinical decision-support systems (CDSS) are being developed with the idea to help clinicians in complex situations (8). Unfortunately, there is no common semantic structure defined, which would facilitate precise and structured documentation and communication of the data representing a breast cancer situation.

To address this issue, we aimed to create a data structure based on radiotherapeutic decision-making criteria used in clinical practice. By using the CDE concept, with such an approach it is potentially possible to create a universal data input layer for any further system of decision-making afterwards, including software systems as well as human evaluators.

## METHODS

### Identifying relevant CDEs used in clinical decision-making for breast cancer patients to undergo radiation therapy

To create and apply the data structure the following steps were conducted. A schematic overview is presented in Fig. 1.

**Fig. 1:**
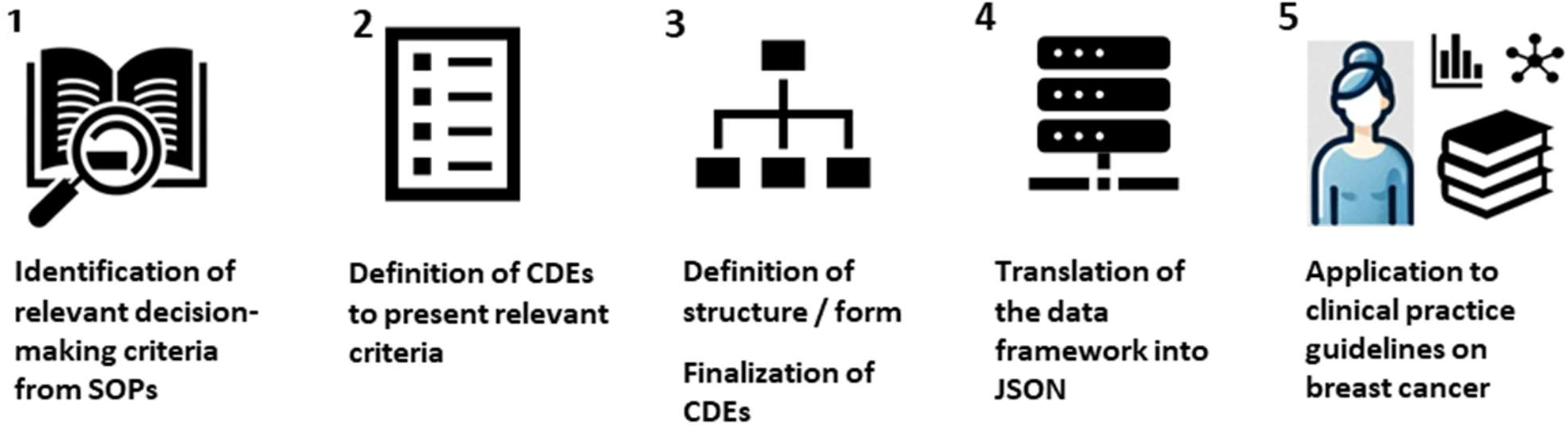
Schematic illustration of the steps used in the methodology for creating and applying the defined CDE-based data structure for radiotherapeutic decision-making in breast cancer.

#### Step 1 - Identification of criteria

The SOPs on radiotherapy in breast cancer patients of the Kantonsspital St. Gallen (Switzerland), which are used for decision-making in clinical practice, were thoroughly analyzed. FD and PMP, who use these SOPs as part of their clinical decision-making, reviewed the documents to identify and formalize criteria involved in different oncological situations in the decision-making process. The physicians discussed the criteria named in the SOPs and confirmed that these are essential for everyday decision-making in the clinical routine. A list of defined criteria was collected.

#### Step 2 - Definition of corresponding CDEs

The criteria were translated into a formalized structure by defining corresponding CDEs. A defined representation of each criterion was created, consisting of one or several “Value List”- or “Number”-CDEs. Regarding “Value List”-CDEs, the list of “Permissible Values” for each CDE were defined, while regarding “Number”-CDEs, corresponding units were defined. The individual CDEs were iteratively revised and refined by FD, MS and NC until consensus was reached and all ambiguities had been resolved. The set of CDEs was confirmed by all three researchers.

#### Step 3 - Definition of structure / form and finalization of CDEs

While CDEs are basically questions that are to be answered for a certain situation, these questions are being asked in a defined setting and regarding a defined concept. As for example, for the concept “breast cancer disease”, a CDE “type of conducted surgery” may be defined. Formulating it as a question, it would mean “What type of surgery was conducted for the breast cancer disease?”.

A “breast cancer disease” may consist of one or multiple “tumor lesions” (e.g., satellite lesions or metastases). In this scenario, a CDE like “tumor size” would make sense on the level of the concept “tumor lesion” but not on the level of “breast cancer disease” (“What size is the tumor lesion?” and not “What size is the breast cancer disease?”). The CDEs were therefore assigned to concepts within the data structure where they should be applied. A hierarchical structure was thereby created. While this structuring is not per se inherent to the CDEs as broadly defined by the NIH, a corresponding delineation is realized with CDE forms (9). It should overall be noted that the structuring of data in the clinical domain using hierarchies or graphs is a well-established approach in other forms of data standards such as e.g., SNOMED (10).

The structuring and finalization was done by FD, MS and NC and again iteratively revised until consensus was reached.

#### Step 4 – Translation of the drafted data framework into machine-readable JSON format

To realize a determined and also computer-readable format, the structure was translated into the JavaScript Object Notation (JSON) format.

A simplified JSON object encompassing the structure was created. Since JSON is also used by the NIH for defining CDEs, our JSON object was created in analogy to this.

Individual oncological situations can then be presented using the provided JSON object by replacing the CDE concept descriptions with the corresponding values of the situation.

#### Step 5 – Application of the data framework to ASTRO practice guidelines for radiotherapeutic decision-making in breast cancer patients

After completion of the data structure, it would be ready to be used for the presentation of individual breast cancer situations and application for radiotherapeutic decision-making as defined within the local SOPs. To investigate to what extent this data structure could also be used for radiotherapeutic decision-making outside of the local environment, it was applied to the clinical practice guidelines for breast cancer of the American Society for Radiation Oncology (ASTRO).

The ASTRO currently (state June 2024) has published six clinical practice guidelines on breast cancer, covering different specific areas (11).

Namely, these are:

- The ASTRO Guideline on Partial Breast Irradiation for Patients With Early-Stage Invasive Breast Cancer or Ductal Carcinoma In Situ, published in 2023 **(PBI-Guideline)** (12)
- The consensus guideline of the American Society of Clinical Oncology (ASCO), the ASTRO and the Society of Surgical Oncology (SSO) on the Management of Hereditary Breast Cancer, published in 2020 **(Hereditary Breast Cancer-Guideline)** (13)
- The ASTRO Evidence-Based Guideline on Radiation Therapy for the Whole Breast, published in 2018 **(WBI-Guideline)** (14)
- The SSO/ASTRO/ASCO Consensus Guideline on Margins for Breast Conserving Surgery with Whole Breast Irradiation in Ductal Carcinoma in Situ (DCIS), published in 2016 **(Margins-DCIS-Guideline)** (15)
- The ASCO/ASTRO/SSO Postmastectomy Radiotherapy Guideline, first published in 2001 and updated in 2016 **(PMRT-Guideline)** (16)
- The SSO/ASTRO Consensus Guideline on Margins for Breast-Conserving Surgery with Whole-Breast Irradiation in Stages I and II Invasive Breast Cancer, published in 2014 **(Margins-BC-Guideline)** (17)

These guidelines and consensus papers contain valuable recommendations for practical radiotherapeutic decision-making in treating breast cancer patients. Similar to the local SOPs, the recommendations also contain various criteria the individual decision is based on.

All the recommendations and mentioned criteria of the six practice guidelines were analyzed in full-text to identify all mentionings of some sort of decision-making criteria in the individual guideline recommendations. For each case it was evaluated if the mentioned decision-making criterion could be displayed using the data structure. For each recommendation, it was checked whether the entire situation with all mentioned criteria could be presented with the data structure (meaning all the mentioned criteria were presentable). The evaluation was done by FD and NC.

## RESULTS

### Criteria and CDEs for radiotherapeutic decision-making in breast cancer patients

In step 1 a total of 31 different criteria that are involved in radiotherapeutic decision-making for breast cancer patients were identified when analyzing the SOP documents. These involved general person-related criteria, criteria regarding tumor location, criteria related to cancer stage, histopathological and genetic criteria as well as criteria about previously conducted oncological therapies.

In step 2 a total of 46 CDEs were defined to present these criteria. These included 36 “Value List”-CDEs and 10 “Number”-CDEs. There were no CDEs of the other data types defined by the NIH (being “Text”, “Date”, “File” and “Externally Defined”). For 18 of the criteria exactly one corresponding CDE was defined. In 12 cases, two CDEs were defined to describe the criterion. For one criterion (=tumor size), four corresponding CDEs were defined.

A basic overview of the criteria and the corresponding CDEs used to describe them is provided in Table 1. More detailed information and description of the CDEs is provided in Appendix 2.

**Table 1:** List of identified criteria for radiotherapeutic decision-making in breast cancer patients together with CDEs defined to represent them.

|  | Name of criterion | Name of CDE | Type of CDE |
| --- | --- | --- | --- |
| General person-related information |  |  |  |
|  | Age | Age | Number |
|  | Menopausal Status | Menopausal Status | Value List |
| Tumor location |  |  |  |
|  | Laterality | Laterality | Value List |
|  | Tumor location | Tumor location | Value List |
| Cancer stage |  |  |  |
|  | cT-Stage | cT-Stage<br>Primary or Recurrent (cT) | Value List<br>Value List |
|  | pT-Stage | pT-Stage<br>Primary or Recurrent (pT) | Value List<br>Value List |
|  | cN-Stage | cN-Stage<br>Primary or Recurrent (cN) | Value List<br>Value List |
|  | pN-Stage | pN-Stage<br>Primary or Recurrent (cN) | Value List<br>Value List |
|  | cM-Stage | cM-Stage<br>Primary or Recurrent (cM) | Value List<br>Value List |
|  | pM-Stage | pM-Stage<br>Primary or Recurrent (pM) | Value List<br>Value List |
|  | Sentinel lymph node status | sentinel lymph node status | Value List |
|  | Lymph node involvement in the mamma interna region | Lymph node involvement in the mamma interna region | Value List |
|  | Invasive carcinoma with associated DCIS | Primary or Recurrent<br>associated DCIS of other tumor lesion | Value List<br>Value List |
| Further histopathological and genetic criteria |  |  |  |
|  | G status | G status | Value List |
|  | L status | L status | Value List |
|  | V status | V status | Value List |
|  | R status | R status | Value List |
|  | Her2-receptor status | Her2-receptor status (IHC)<br>Her2-receptor status (positive/negative) | Value List<br>Value List |
|  | Histological subtype | Histological subtype | Value List |
|  | Extracapsular extension of a lymph node metastasis | Extracapsular extension of a lymph node metastasis | Value List |
|  | BRCA status | BRCA1-Status<br>BRCA2-Status | Value List<br>Value List |
|  | Minimal resection margin | Minimal resection margin | Number |
|  | Tumor size | Diameter1<br>Diameter2<br>Diameter3<br>Modality of Assessment | Number<br>Number<br>Number<br>ValueList |
|  | Number of positive resected lymph nodes | Number of positive resected lymph nodes | Number |
|  | Number of resected lymph nodes | Number of resected lymph nodes | Number |
|  | Estrogen receptor expression | ER status (%)<br>ER status (positive/negative) | Number<br>Value List |
|  | Progesterone receptor expression | PR status (%)<br>PR status (positive/negative) | Number<br>Value List |
|  | Ki-67 | Ki-67 status (%) | Number |
| Previously conducted oncological therapies |  |  |  |
|  | Conduction of neoadjuvant chemotherapy | Conduction of neoadjuvant chemotherapy | Value List |
|  | Post-resection performed | Post-resection performed | Value List |
|  | Previous surgery | Previous surgery conducted<br>Type of previously conducted surgery | Value List<br>Value List |

### Conceptualization of the data structure

In step 3 a hierarchy with presentation of the CDEs in an object-oriented structure was created. Four main classes, representing four conceptual levels, within which CDEs are used, were defined, namely “Patient”, “BreastCancerDisease”, “TNM” (= Classification system to stage the cancer situation based on Tumor, Lymph-Node and Metastasis) and “TumorLesion” (Fig. 2).

**Fig. 2:**
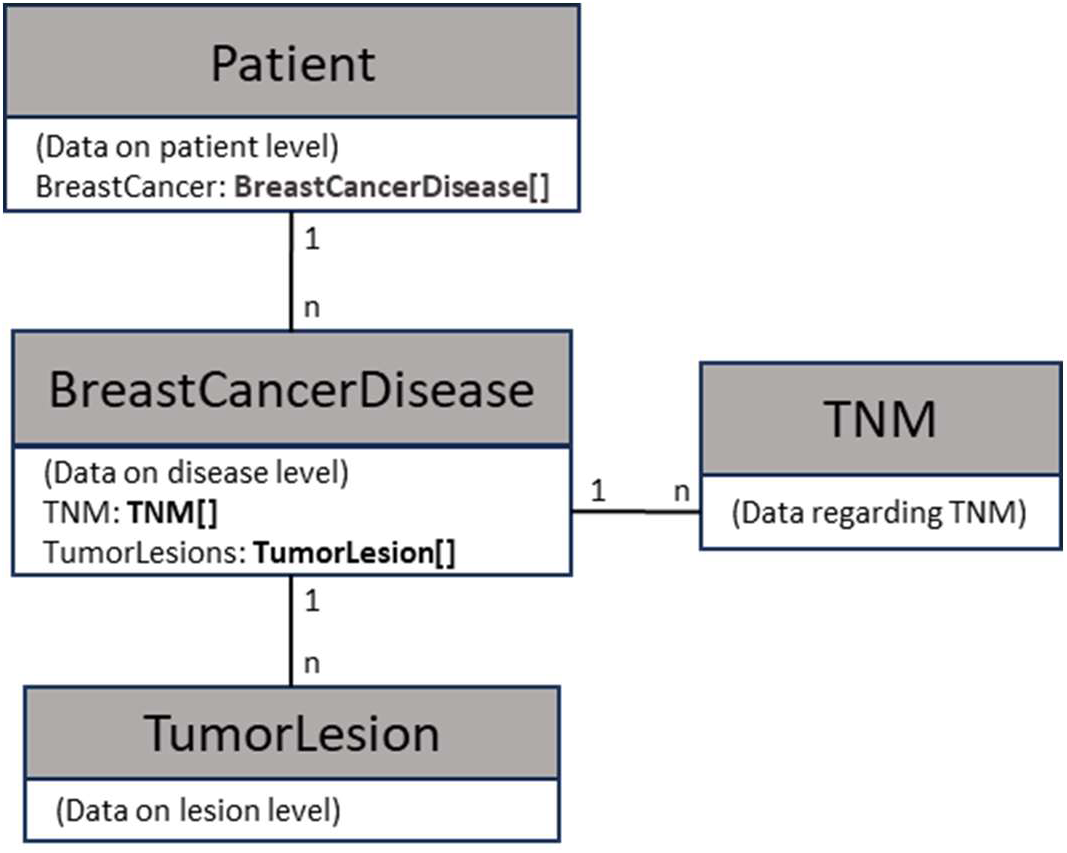
Schematic illustration of the four main concepts (=classes) and relationships used in the data structure.

The class “Patient” has the attribute BreastCancer, which is an array of objects of type “BreastCancerDisease”. “BreastCancerDisease” has the attributes TNM (Array of type TNM) and TumorLesions (Array of type TumorLesion). The rationale of this structuring is the following: A patient can have zero, one or more than one independent breast cancer diseases. A breast cancer disease consists of one or several tumor lesions (including recurrent lesion, associated DCIS, satellite lesion or metastases; lymph node metastases are handled separately). Each breast cancer disease can be staged according to TNM (it should be noted that there exist several TNM classification systems, most notably according to UICC and to AJCC; it should be clearly defined, which system is used – in our study we used the UICC system; see also Appendix 2). There can be several TNM stagings of a disease, including differences regarding “clinical” and “pathological” staging as well as stagings happening at different time points, that may all be relevant for clinical decision-making.

Further subclasses for organizing and structuring of the CDEs were defined – an example for “TumorLesion” with the subclasses “location of tumor lesion”, “general data about tumor lesion” and “tumor size” is provided in Fig. 3.

**Fig. 3:**
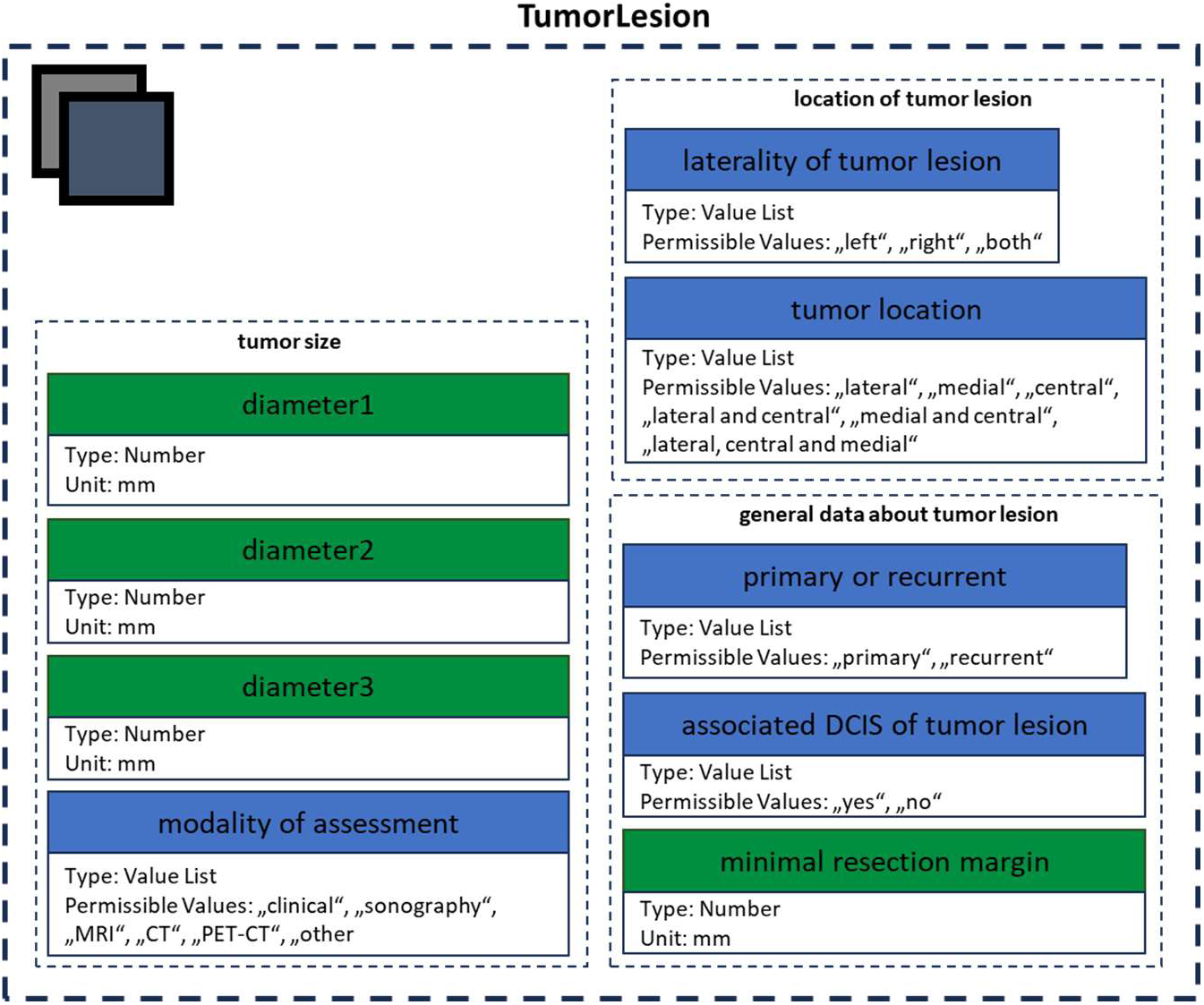
Structure of the class “TumorLesion”, which contains the subclasses “location of tumor lesion”, “general data about tumor lesion” and “tumor size”. “Value List”-CDEs are presented in blue, “Number”-CDEs in green.

The overall structure as a diagram is provided in Appendix 1 while a list with the parameters of the CDEs and further descriptions is provided in Appendix 2.

All the subclasses have a 1:1 relationship between parent class and subclass. While they are helpful for improved overview when organizing the CDEs, they are not necessary from a semantical point of view and the CDEs could also be defined as direct attributes in the main classes.

In step 4 a corresponding JSON format encompassing the three main classes, subclasses and CDEs was created. The final structure that can be applied to various breast cancer situations, is provided in Appendix 3.

### Application of the data framework to ASTRO practice guidelines

In step 5 the six mentioned clinical practice guidelines for breast cancer published by the ASTRO were analyzed in full-text. A total of 94 recommendations were identified across the guidelines with 90 recommendations mentioning some sort of decision-making criteria 216 times. In 151 cases (70.0 %), the criterion could be presented using the data structure, while in the other 65 cases the required semantic information could not fully be described with the data structure. For 52 of the guideline recommendations (not including the four recommendations that did not mention any decision-making criteria), all mentioned criteria of the recommendation were fully presentable using the data structure (57.7%).

An example of a fully presentable recommendation as well as one example of a not presentable recommendation is illustrated in Fig. 4.

**Fig. 4:**
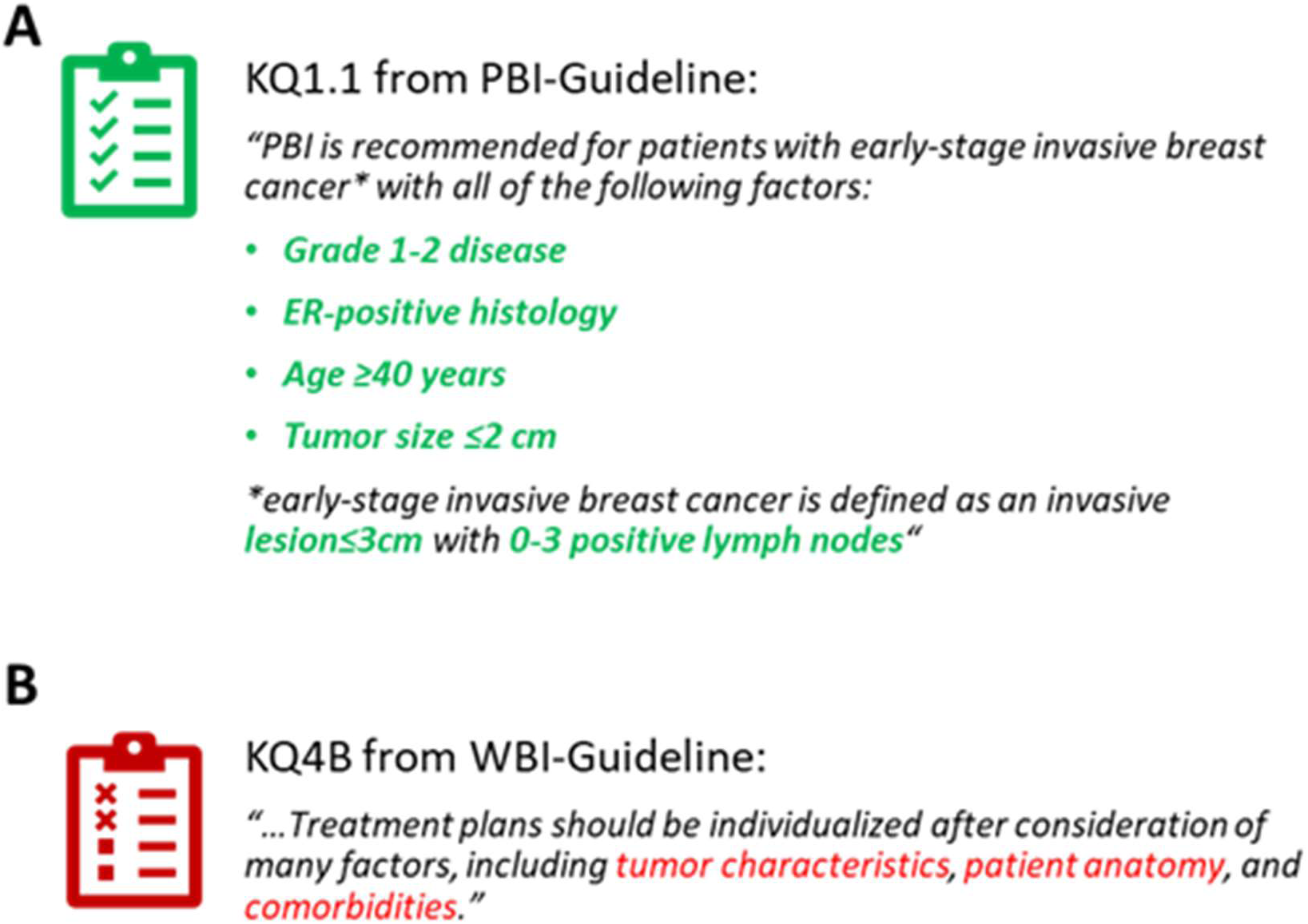
Example of two recommendations from the guidelines that can either be (A) fully presented using the data structure with all mentioned decision-making criteria implemented in the CDE-based data structure or (B) not presentable using the data structure since some of the relevant information involved in the decision-making is not implemented.

The portion of presentable criteria and the amount of guideline recommendations that could be fully presented varied among the six guidelines (see Table 2).

**Table 2:** Amount of presentable recommendations and criteria in the six clinical practice guidelines of the ASTRO.

| Practice Guideline | Presentable criteria / total number of criteria | Presentable recommendations / total number of recommendations |
| --- | --- | --- |
| PBI-Guideline | 53/56 (94.6%) | 17/20 (85.0%) |
| Hereditary Breast Cancer-Guideline | 25/52 (48.1%) | 8/22 (36.4%) |
| WBI-Guideline | 35/56 (62.5%) | 12/27 (44.4%) |
| Margins for DCIS-Guideline | 10/11 (90.1%) | 7/8 (87.5%) |
| PMRT-Guideline | 18/26 (69.2%) | 4/6 (66.7%) |
| Margins for Breast Cancer-Guideline | 10/15 (66.7%) | 4/7 (57.1%) |

The data with the individual criteria mentioned in the recommendations of the six guidelines and the CDEs of the data structured used to present them is provided in Appendix 4.

## DISCUSSION

### Providing semantic information using CDEs

CDEs are a powerful yet simple concept that focus on the meaning of medical information (18). If used within an appropriate framework, CDEs can be used to clearly present the semantic data of a medical situation. Despite radiation oncology being a technical and data-driven discipline, the literature on CDEs and structured documentation in radiation oncology is scarce (5). While there have been efforts made regarding definition and clarification of CDEs in the domain of breast cancer for some research registries (19), there is no broader framework for structured documentation of breast cancer data that would be used commonly among different stakeholders (particularly not in clinical practice). At the same time, there are obviously corresponding semantic concepts and data structures being used within the logic of clinical software and forms/documents (5), (20).

### Using CDEs for data collection in clinical practice and for real-world data

Even though developed for standardized data collection within the setting of clinical trials, CDEs are increasingly being used for data management in daily clinical life (21). In practice, the concept of CDEs can be applied to various media for documentation of medical data, including case report forms (CRF), electronic health records (EHR), clinical information systems (CIS) or analog document forms (22), (23). This idea of using CDEs for documentation in daily clinical life becomes of additional interest regarding the increasing interest in RWD for addressing research questions while supporting daily healthcare (21).

CDEs provide a common language for enabling structural and semantic interoperability. They allow for the alignment of data from various sources like EHR, healthcare claims, and patient-generated data streams. By expressing CDEs in machine-computable formats, data can be mapped, transformed, and combined across disparate sources (24).

### CDE-based structuring of data for radiotherapeutic decision-making on the level of a local hospital

The implementation of a CDE-based system for data collection in a local hospital setting requires a considerable amount of planning, training, and integration with existing concepts, standards and healthcare IT systems. However, the payoff in terms of improved data quality, enhanced monitoring of cancer situations, and streamlined clinical workflows could be substantial (25).

In the dynamic and nuanced field of radiotherapy for breast cancer patients, the utilization of CDEs marks a transformative shift towards personalized and evidence-based treatment. By implementing a CDE-based framework, healthcare professionals could access a consolidated view of a patient’s comprehensive medical history and current health status, enabling the formulation of a tailored radiotherapeutic strategy.

### Leveraging data exchange up to the level of clinical practice guidelines

While the data structure in our work was created as a first concept to formalize decision-making criteria used on the level of a local hospital, we have seen that a considerable amount of the criteria mentioned in international clinical practice guidelines can also be presented using this structure (ranging from 48.1 to 94.6 %; see Table 2). This may not be surprising, since local SOPs should align with the recommendations of established clinical practice guidelines. One may assume that the relevant decision-making criteria are shared on a broader level among different facilities. However, it has been repeatedly shown for many examples in oncological decision-making, that this assumption is clearly wrong (26), (27), (7). As we had also recently seen in a document analysis study that aimed to find shared CDEs among radiotherapy departments when ordering a planning CT, the majority of data cannot be exchanged (23). Overall, more often than not, exchange of standardized data is very limited among different facilities. Neither the local SOPs, nor the six ASTRO guidelines were developed with a focus on interoperability or clear definition and formalization of criteria and concepts. It is not surprising that e.g., for the Hereditary Breast Cancer Guideline, the applicability was limited with presentable criteria at only 48.1% (Table 2). Hereditary breast cancer is a special situation that was not fully addressed in the data structure based on the local SOPs. Nevertheless, the fact that overall in a majority of 70.0% of cases a mentioned criterion in the guidelines was representable, confirms the usage of CDEs for promoting interoperability.

### A shared CDE-based data structure

The data structure could be extended to also cover more of the relevant decision-making criteria mentioned in the ASTRO guidelines to be further applicable. Adjusting the data structure for implementation of international guidelines should be based on broader consensus including clinical experts involved in guideline creation. As the ISROI will continue to work on the structuring of radiotherapeutic data using CDEs, this will be the subject of future initiatives. In any case, both the medical and the informatics perspective need to be kept in mind to clearly define concepts, criteria and CDEs.

One major advantage of using a CDE-based data structure is that it could be used in a modular way without giving up the overall structure itself. For example, one could create an additional subclass for the information regarding hereditary breast cancer to cover the related information. This subclass could be implemented in a more comprehensive version of the data structure and only be used if relevant for a specific question in a certain guideline or local SOP.

If a functioning, well thought out data structure is in place, CDEs could also empower downstream systems like integrated CDSS or clinical algorithms to deliver more accurate, timely, and personalized medical advice and treatment recommendations (28). Since one major goal of CDEs is the promotion of standardization and interoperability, they should not remain merely abstract concepts but be applied in clinical practice at the local hospital level.

If diverse stakeholders were to adopt a common CDE-based data framework, it would enable local hospitals to engage in collaborative decision-making and enhance standardization and data sharing across institutions. Although such a framework has not yet been realized, its implementation is essential for facilitating data exchange among various institutions and expanding the use of IT and AI solutions on a larger scale.

### Limitations, challenges and possible solutions

Even though the CDE-based data structure developed in this work allows for clear documentation and communication of medical data, the approach itself has some limitations. Theoretically, if the values of all the CDEs of a breast cancer situation are known, any such situation could be unambiguously presented in the proposed JSON format. However, in clinical practice, ambiguities, uncertainties and contradictory data are common. To use the framework on real data encompassing such ambiguities, uncertainties and contradictions, one could define a clear methodology to assign CDE values to real world situations. However, for versatile situations like breast cancer, even very sophisticated methods can not cover every possible situation that may need to be addressed during such value assignment (29). Yet, this is more a fundamental problem of suboptimal and ambiguous data than of the structuring approach itself.

Nevertheless, the structure in its current form as it is based on the original definition of CDEs by the NIH is not all-encompassing. One issue is the fact that the value of any data element is assessed at one certain time point, which is not addressed in the data structure. An obvious example is the CDE “Age”, which changes as the patient gets older. The values are therefore not necessarily static but may have a dynamic that is not implemented currently. One possible solution would be to extend the concept of CDEs within the framework and define the values with an additional (optional) timestamp. In situations where multiple data points for a CDE are provided, the corresponding attribute in the JSON object could be presented with an array of objects containing the value and (if known) the time stamp. A similar idea of such an array-format is in the current structure only provided for the concept “TNM”, as it was considered a relevant dynamic factor in the analyzed SOPs.

A further interesting idea to extend the possibilities of CDE-based data documentation was proposed by Kim et al. (30). They introduced so-called “composite relationships” and defined the constraints “operated”, “required”, “dependent” and “ordered” in a proposed extension of the CDE concept. Defining such relationships, it is possible to establish a broader semantic framework.

However, even if very complex systems were used, it would be very challenging to depict not only the data of broader, well-established concepts, but also detailed information and nuances of a medical situation. If CDEs are to be used for facilitating interoperability, they need to be implemented on a local level as well as on a broader abstract level. To address individual needs, also the criteria used only by a few actors (so-called insular criteria (31)) would need to be addressed since they may influence local decision-making. Overall, the relevant criteria and corresponding CDEs need clear definitions. The reason why many criteria mentioned in the ASTRO guidelines could not be presented with the presented data structure is due to unclear/missing definitions. Criteria like “tumor characteristics”, “patient anatomy” or “comorbidities” (see Fig. 4B) are certainly relevant, but are not clearly defined within the analyzed guidelines. As a result, these criteria remain ambiguous. Taking the effort of presenting criteria with structures based on CDEs actually formalizes these criteria and thereby promotes standardization and interoperability while reducing ambiguities. Clearly defining rather abstract criteria may take a considerable amount of effort, but is possible (as for example done for “comorbidities” by the National Cancer Institute (NCI) on the US population (32)). It has to be acknowledged however, that not every possible criterion can be formalized using the CDE concept (e.g., a criterion like “high risk of complications“). Presenting medical information with CDEs resembles the creation of a synoptic report, which gives a concise and clear overview, but a priori is a simplification.

The presented data structure in this work is not an all-encompassing framework that could be used on a broader inter-institutional level to collect all relevant information for radiotherapeutic decision-making. Current definitions are also not truly unambiguous, so that all possible situations and questions can be answered (see also further information on the CDEs in Appendix 2). The CDEs, classes and subclasses of the data structure will need to be extended, refined and aligned with existing medical data standards in the future. However, it may be a starting point originating from real world clinical decision-making. It can build a basis for further initiatives conducted by the ISROI and other stakeholders to promote structuring and formalization of relevant data and criteria used in clinical practice for radiation therapy and general oncology.

The study itself has some limitations. While multiple researchers designed and iteratively refined the framework both from a medical and a data science perspective, the used methodology is to some extent subjective. Furthermore, no systematic search alignment with existing CDE resources has been conducted. More sophisticated and elaborate methods like Delphi rounds among independent researchers will be needed, if a standardized data structure is to be established for documentation and exchange of data across different stakeholders on an inter-institutional level.

## CONCLUSIONS

The development of a CDE-based data structure for radiotherapy in breast cancer represents an advance in organizing medical data for clinical use. Relevant decision-making criteria can be identified and translated into CDEs for organizing medical data in a structured and clear way. This allows for direct connection of information with other data systems such as software based IT/AI applications. At the same time, the application of the data framework to clinical practice guidelines showcases the CDEs’ utility in clarifying data and facilitating interoperability. The work emphasizes the importance of adopting structured data practices in radiation oncology to enable data-driven, personalized patient care.

## Data Availability

All the data obtained in the study (created CDEs and structures) are provided in this work or its Supplementary Material. The six ASTRO guidelines used in the study have previously been published with the publications listed in the References.

## List of abbreviations

AI: artificial intelligence
AJCC: American Joint Committee on Cancer
ASCO: American Society of Clinical Oncology
ASTRO: American Society for Radiation Oncology
CDE: Common Data Element
CDSS: Clinical Decision Support System
CIS: Clinical Information System
CRF: Case Report form
EHR: Electronic Health Record
ISROI: International Society for Radiation Oncology
IT: information technology
JSON: JavaScript Object Notation
Hereditary Breast Cancer-Guildeline: ASTRO/ASCO/SSO Consensus guideline on the Management of Hereditary Breast Cancer
Margins-BC-Guideline: SSO/ASTRO Consensus Guideline on Margins for Breast-Conserving Surgery with Whole-Breast Irradiation in Stages I and II Invasive Breast Cancer
Margins-DCIS-Guideline: SSO/ASTRO/ASCO Consensus Guideline on Margins for Breast Conserving Surgery with Whole Breast Irradiation in DCIS
NCI: National Cancer Institute
NIH: National Institutes of Health
NLP: Natural Language Processing
PBI-Guideline: ASTRO Guideline on Partial Breast Irradiation for Patients With Early-Stage Invasive Breast Cancer or Ductal Carcinoma In Situ
PMRT-Guideline: ASCO/ASTRO/SSO Postmastectomy Radiotherapy Guideline
RO: Radiation Oncology
RWD: real-world data
SSO: Society of Surgical Oncology
SOP: standard operating procedure
UICC: Union for International Cancer Control
WBI-Guideline: ASTRO Evidence-Based Guideline on Radiation Therapy for the Whole Breast

## Ethics approval and consent to participate

Not applicable. This study did not involve any human participants, biological material, or personal data. Consequently, it does not fall under the jurisdiction of the Human Research Act, and no ethics committee approval was required (https://www.fedlex.admin.ch/eli/cc/2013/617/de).

## Availability of data and materials

All the data obtained in the study (created CDEs and structures) are provided in this work or its Supplementary Material. The six ASTRO guidelines have previously been published with the publications listed in the References (12), (13), (14), (15), (16), (17).

## Authors’ contribution

Conceptualization, F.D, M. Schmalfuss., J. Z., J. H., N. C.; methodology, F.D., M. Schmalfuss., M. Schmerder; formal analysis, F.D., M. Schmalfuss, R. G.; writing—original draft preparation, F.D., M. Schmalfuss.; writing—review and editing, all authors; supervision, PM. P., N. C.; project administration, F.D., N.C.

## Acknowledgements

Not applicable.

