## Supplementary figures and images for "A CDE-based data structure for radiotherapeutic decision-making in breast cancer"

### Appendix1

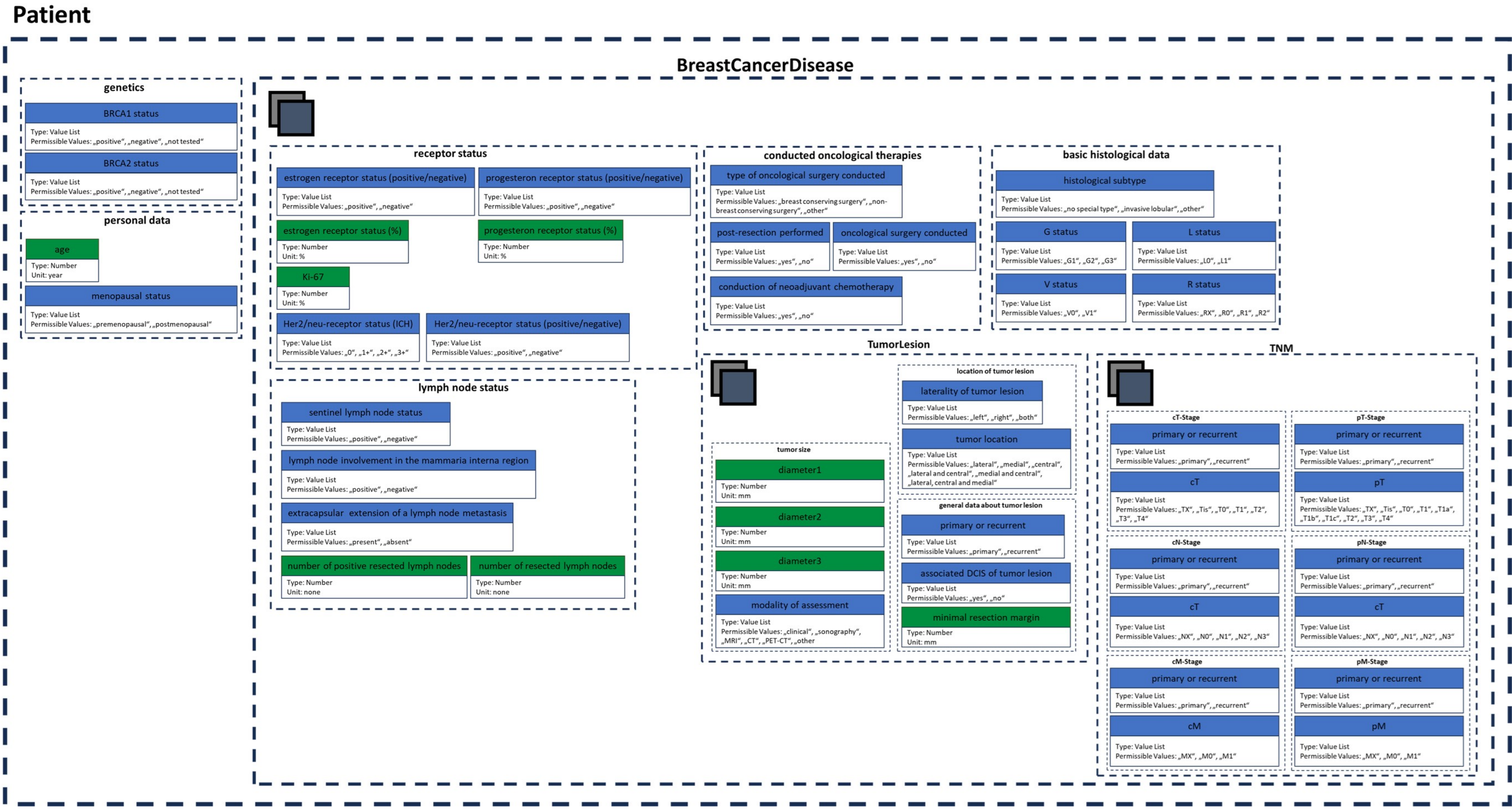
