## Appendix2 for "A CDE-based data structure for radiotherapeutic decision-making in breast cancer"

**Appendix 3: CDEs used in the classes of the data structure**

**Class ‘Genetics’**

- **BRCA1 status:**
  - Type: Value List
  - Permissible Values: ‘positive’, ‘negative’, ‘not tested’
  - Description: Determines whether the patient has a mutation in the BRCA1 gene associated with a higher risk for breast cancer.
  - Notes: It should be noted that determining the BRCA1 status can be ambiguous. The interpretation of genetic testing results may vary based on the specific mutations identified and the techniques used in the testing process. Variants of uncertain significance may also be identified, which are not clearly classified as positive or negative. Furthermore, discrepancies in testing methods or laboratory standards can lead to different interpretations of the same genetic data.
- **BRCA2 status:**
  - Type: Value List
  - Permissible Values: ‘positive’, ‘negative’, ‘not tested’
  - Description: Determines whether the patient has a mutation in the BRCA2 gene associated with a higher risk for breast cancer.
  - Notes: It should be noted that determining the BRCA2 status can be ambiguous. The interpretation of genetic testing results may vary based on the specific mutations identified and the techniques used in the testing process. Variants of uncertain significance may also be identified, which are not clearly classified as positive or negative. Furthermore, discrepancies in testing methods or laboratory standards can lead to different interpretations of the same genetic data.

**Class ‘Personal Data’**

- **Age:**
  - Type: Number
  - Unit: year
  - Description: The current age (meaning at the time of the application of the data structure) of the patient in years.
  - Notes: Age is of course a dynamic factor that changes over time, as an individual gets older. Therefore the “current” age, meaning the time point the data structure is used, is the correct one. Obviously, the age could be inferred from other data, like e.g., birthdate and current date. However, addressing inference of data is not the purpose of this work, since it requires exact formulations even for seemingly simple inference (e.g., for “Age” one would need to define how to handle special cases like individuals born on 29^th^ of February or differences in time zones etc.).
- **Menopausal Status:**
  - Type: Value List
  - Permissible Values: ‘premenopausal’, ‘postmenopausal’
  - Description: Data on whether the patient is premenopausal or postmenopausal.
  - Notes: It should be noted, that determination of the Menopausal status may be challenging, since a clear clinical diagnosis is not always possible.

**Class ‘Receptor status’**

- **Estrogen receptor status (%):**
  - Type: Number
  - Unit: %
  - Description: The %-value of the estrogen receptor status of the breast cancer disease.
  - Notes: It should be noted that cancer consists of a heterogeneous collection of tumor cells with heterogeneous and changing expression of the estrogen receptor on the surface of cells. The exact portion that express the estrogen receptor is never known and can only be estimated from bioptic or histological samples that are also only analyzed in part with the result being furthermore dependent on the pathological methods used. In clinical practice, the true value cannot be known, but has to be estimated.
- **Estrogen receptor status (positive/negative):**
  - Type: Value List
  - Permissible Values: ‘positive’, ‘negative’
  - Description: A binary value for the estrogen receptor status of the breast cancer disease (positive or negative).
  - Notes: Whether or not a breast cancer disease is considered to be estrogen receptor positive or negative is of course subjective. Usually, a threshold of the percentage of positive tumor cells is defined to separate positive from negative breast cancer diseases. However, since the percentage value is only estimated (see above), inconsistent and depending on the method of measurement, there may be certain situations where a clear determination may not be possible in clinical practice.
- **Progesterone receptor status (%):**
  - Type: Number
  - Unit: %
  - Description: The %-value of the progesterone receptor status of the breast cancer disease.
  - Notes: It should be noted that cancer consists of a heterogeneous collection of tumor cells with heterogeneous and changing expression of the progesterone receptor on the surface of cells. The exact amount that express the progesterone receptor is never known and can only be estimated from bioptic or histological samples that are also only analyzed in part with the result being furthermore dependent on the pathological methods used. In clinical practice, the true value cannot be known, but has to be estimated.
- **Progesterone receptor status (positive/negative):**
  - Type: Value List
  - Permissible Values: ‘positive’, ‘negative’
  - Description: A binary value for the progesterone receptor status of the breast cancer disease (positive or negative).
  - Notes: Whether or not a breast cancer disease is considered to be progesterone receptor positive or negative is of course subjective. Usually, a threshold of the percentage of positive tumor cells is defined to separate positive from negative breast cancer diseases. However, since the percentage value is only estimated (see above), inconsistent and depending on the method of measurement, there may be certain situations where a clear determination may not be possible in clinical practice.
- **Her2/neu-receptor status (IHC):**
  - Type: Value List
  - Permissible Values: ‘0’, ‘1+’, ‘2+’, ‘3+’
  - Description: Value for the immunohistochemistry (IHC) status of the receptor of the Her2/neu for the breast cancer disease.
  - Notes: The Her2/neu-receptor status determined by immunohistochemistry (IHC) can sometimes be ambiguous, especially for results classified as '2+' which are considered equivocal. In such cases, further testing using the Fluorescence In Situ Hybridization (FISH) method is often recommended to clarify the Her2 status. Additionally, discrepancies in IHC results may arise due to variations in laboratory procedures, antibody specificity, and interpretation criteria.
- **Her2/neu-receptor status (positive/negative):**
  - Type: Value List
  - Permissible Values: ‘positive’, ‘negative’
  - Description: A binary value for the Her2/neu receptor status of the breast cancer disease (positive or negative). Even though it is recommended to use IHC class instead of positive/negative, the latter one is still used and can contain some information, which can be presented with this CDE.
  - Notes: The binary classification of Her2/neu-receptor status as 'positive' or 'negative' simplifies the reporting but can sometimes mask underlying complexities and nuances present in the IHC scoring system. Usually, ‘positive' results correspond to an IHC score of '3+', while a 'negative' result typically corresponds to an IHC score of '0' or '1+'. However, the '2+' category requires additional testing to resolve its equivocal nature, which can affect the binary classification. As mentioned, the presented data structure in this work does not include inference/interpretation of data with the structure.
- **Ki-67:**
  - Type: Number
  - Unit: %
  - Description: The %-value of the Ki-67 status of the breast cancer disease.
  - Notes: It should be noted that cancer consists of a heterogeneous collection of tumor cells with heterogeneous and changing expression of Ki-67. In practice, the value is of course only measured for a given tumor sample. Factors such as differences in staining techniques, the specific antibody used, and the threshold chosen for positivity can influence the Ki-67 value.

**Class ‘Conducted oncological therapies’:**

- **Oncological surgery conducted:**
  - Type: Value List
  - Permissible Values: ‘yes’, ‘no’
  - Description: A binary value about whether an oncological surgery has been conducted.
  - Notes: The binary classification of whether oncological surgery has been conducted simplifies documentation but may not capture the full scope and context of the surgical intervention. It may include various types of oncological surgeries, ranging from minimally invasive procedures to extensive resections.
- **Type of oncological surgery conducted:**
  - Type: Value List
  - Permissible Values: ‘breast conserving surgery’, ‘not-breast conserving surgery’, ‘other’
  - Description: Value about what general type (breast conserving, not-breast conserving, other) of oncological surgery has been conducted.
  - Notes: The classification of the type of oncological surgery provides a broad categorization of the surgical approach taken. However, these categories may not fully encapsulate the specific nature or complexity of the surgical procedures performed. For instance, "breast conserving surgery" can range from a simple lumpectomy to more complex oncoplastic techniques, while "not-breast conserving surgery" includes various forms of mastectomy with or without reconstruction. The category "other" serves as a catch-all for procedures that do not fit neatly into the first two categories, but it lacks specificity.
- **Post-resection performed:**
  - Type: Value List
  - Permissible Values: ‘yes’, ‘no’
  - Description: Binary value about whether a ‘post-resection’, meaning a second resection after first initial resection has been conducted.
  - Notes: This binary value alone does not provide information on the reasons for the post-resection, the extent of the second surgery, or its outcomes. Additional context, such as pathology reports, surgical notes, and the timing between the initial and second resections, might be relevant for a comprehensive understanding of the patient's surgical history.

**Conduction of neoadjuvant chemotherapy:**

- Type: Value List
- Permissible Values: ‘yes’, ‘no’
- Description: Binary value about whether a neoadjuvant chemotherapy was given before conduction of the oncological surgery.
- Notes: This binary value does not convey information about the specific chemotherapy regimen used, the duration of treatment, the patient's response to the therapy, or any side effects experienced.

**Class ‘Basic histological data’:**

- **Histological subtype:**
  - Type: Value List
  - Permissible Values: ‘no special type’, ‘invasive lobular’, ‘other’
  - Description: Data about the histological subtype of breast cancer.
  - Notes: The permissible values categorize the cancer into general types: 'no special type' (NST), which is the most common subtype and includes ductal carcinoma, 'invasive lobular', and 'other', which encompasses less common subtypes. While this classification is helpful, it may not capture the full spectrum of histological diversity within breast cancer. Some subtypes included in 'other' might have distinct biological behaviors and treatment responses. Additionally, accurate classification relies on high-quality tissue samples and expert pathological assessment. Therefore, detailed pathology reports, including specific histological features and any relevant molecular finding may include additional relevant information.
- **G status:**
  - Type: Value List
  - Permissible Values: ‘G1’, ‘G2’, ‘G3’
  - Description: Data about the grading (G status) of the cancer disease.
  - Notes: The G status, or histological grade, of cancer indicates how much the tumor cells differ from normal cells and how quickly the tumor is likely to grow and spread. It is categorized into three grades: G1 (well-differentiated), G2 (moderately differentiated), and G3 (poorly differentiated). This grading provides crucial information about the aggressiveness of the tumor. However, the interpretation of G status can sometimes be subjective, depending on the pathologist's experience and the criteria used for grading. Moreover, intra-tumoral heterogeneity, where different parts of the tumor might have different grades, can also complicate the assessment.
- **L status:**
  - Type: Value List
  - Permissible Values: ‘L0’, ‘L1’
  - Description: Binary value about the lymphatic invasion (L status) of the cancer disease.
  - Notes: The L status provides information on the presence or absence of lymphatic invasion by the cancer. 'L0' indicates no lymphatic invasion, while 'L1' indicates that cancer cells have invaded the lymphatic vessels. Assessing lymphatic invasion can be challenging and may be subject to variability due to differences in tissue sampling, staining techniques, and pathological interpretation. In some cases, lymphatic invasion might be focal and not uniformly distributed across the tumor, leading to potential underestimation or overestimation of the L status.
- **V status:**
  - Type: Value List
  - Permissible Values: ‘V0’, ‘V1’
  - Description: Binary value about the vascular invasion (V status) of the cancer disease.
  - Notes: The V status indicates the presence or absence of cancer cells within blood vessels, with 'V0' representing no vascular invasion and 'V1' indicating vascular invasion. Identifying vascular invasion can be challenging and may vary depending on the quality of the tissue sample, the staining techniques used, and the pathologist's expertise. Vascular invasion might be focal and not uniformly present throughout the tumor, leading to potential variability in assessment.
- **R status:**
  - Type: Value List
  - Permissible Values: ‘RX’, ‘R0’, ‘R1’, ‘R2’
  - Description: Value about the resection status (R status) of the cancer disease.
  - Notes: The R status provides information on the status of the surgical resection margins after tumor removal. The classifications are as follows: 'RX' indicates that the resection margin cannot be assessed, 'R0' indicates no residual tumor (clean margins), 'R1' indicates microscopic residual tumor, and 'R2' indicates macroscopic residual tumor. This evaluation can be influenced by the quality of the surgical specimen, the thoroughness of the pathological examination, and the techniques used to examine the margins. Additionally, variability in surgical techniques and the anatomical complexity of the tumor location can affect the R status assessment.

**Class ‘lymph node status’:**

- **sentinel lymph node status:**
  - Type: Value List
  - Permissible Values: ‘positive’, ‘negative’
  - Description: Binary value about whether there is a positive sentinel lymph node.
  - Notes: A 'positive' status indicates that cancer cells have been found in the sentinel lymph node. A 'negative' status means no cancer cells were detected in the sentinel lymph node. The accuracy of this assessment can be influenced by the thoroughness of the sentinel lymph node biopsy, the pathologist's expertise, and the methods used for detecting cancer cells (e.g., standard histopathology vs. immunohistochemistry). False negatives can occur if micrometastases or isolated tumor cells are missed.
- **lymph node involvement in the mammaria interna region:**
  - Type: Value List
  - Permissible Values: ‘positive’, ‘negative’
  - Description: Binary value about whether or not a lymph node in the mammaria interna region is positive.
  - Notes: A 'positive' result indicates the presence of cancer cells in the internal mammary lymph nodes. A 'negative' result indicates no cancer cells were found in these lymph nodes. The accuracy of detecting lymph node involvement in this region can be challenging due to the anatomical location and accessibility of these nodes. Imaging techniques and surgical expertise play a considerable role in accurately identifying and sampling these lymph nodes.
- **Number of positive resected lymph nodes:**
  - Type: Number
  - Unit: none
  - Description: The total number of lymph nodes that were resected and had confirmed cancer of the breast cancer disease.
  - Notes: The number of positive resected lymph nodes can be influenced by the extent of lymph node dissection performed during surgery, the thoroughness of pathological examination, and the techniques used to detect cancer cells within the nodes. The value is of course dependent on the accuracy of detecting tumor cells with pathological methods.
- **Number of totally resected lymph nodes:**
  - Type: Number
  - Unit: none
  - Description: The total number of lymph nodes that were resected as part of the breast cancer treatment, independent of whether or not they had confirmed cancer.
  - Notes: The total number of lymph nodes resected can vary based on the surgical technique, the surgeon's expertise, and the anatomical variability among patients.
- **Extracapsular extension of a lymph node metastasis**
  - Type: Value List
  - Permissible Values: ‘present’, ‘absent’
  - Description: Binary value about whether or not any positive lymph node had cancer with extracapsular extension.
  - Notes: The presence of extracapsular extension (ECE) in a lymph node metastasis indicates that cancer cells have spread beyond the lymph node capsule into the surrounding tissues. The assessment of ECE can be subject to variability depending on the quality of the pathological examination and the criteria used to define ECE.

**Class ‘TNM’:**

- **PrimaryOrRecurrect (for all subclasses)**
  - Type: Value List
  - Permissible Values: ‘primary’, ‘recurrent’
  - Description: Binary value about whether the staging according to TNM (UICC) is in a primary or recurrent situation of the breast cancer disease
  - Notes: A 'primary' classification indicates the initial diagnosis and staging of breast cancer, while 'recurrent' refers to the return of cancer after treatment.
- **cT**
  - Type: Value List
  - Permissible Values: ‘TX’, ‘T0’, ‘Tis’, ‘T1’, ‘T2’, ‘T3’, ‘T4’
  - Description: CDE about the clinical T stage of the breast cancer situation (according to UICC).
  - Notes: It is important to note that there are different versions of the UICC staging system, and the criteria for each T stage might vary slightly between versions. Accurate assessment of the clinical T stage requires a thorough clinical examination, imaging studies, and sometimes pathological assessment to determine the exact size and extent of the tumor.
- **pT**
  - Type: Value List
  - Permissible Values: ‘TX’, ‘T0’, ‘Tis’, ‘T1’, ‘T1a’, ‘T1b’, ‘T1c’, ‘T2’, ‘T3’, ‘T4’
  - Description: CDE about the pathological T stage of the breast cancer situation (according to UICC).
  - Notes: It is important to note that there are different versions of the UICC staging system, and the criteria for each T stage might vary slightly between versions. Accurate assessment of the pathological T stage requires thorough pathological examination, including the size and extent of the tumor and its relationship to surrounding structures.
- **cN**
  - Type: Value List
  - Permissible Values: ‘NX’, ‘N0’, ‘N1’, ‘N2’, ‘N3’
  - Description: CDE about the clinical N stage of the breast cancer situation (according to UICC).
  - Notes: It is important to note that different versions of the UICC staging system exist, and the criteria for each cN stage may vary slightly between versions. Therefore, specifying the version of the UICC staging system used when documenting the clinical N stage is crucial for ensuring consistency and accuracy in staging. Accurate assessment of the cN stage requires a thorough clinical examination, imaging studies (such as ultrasound, CT, or MRI), and sometimes biopsy or other pathological confirmation.
- **pN**
  - Type: Value List
  - Permissible Values: ‘NX’, ‘N0’, ‘N1’, ‘N2’, ‘N3’
  - Description: CDE about the pathological N stage of the breast cancer situation (according to UICC).
  - Notes: It is important to note that different versions of the UICC staging system exist, and the criteria for each pN stage may vary slightly between versions. Accurate determination of the pN stage requires thorough pathological examination, including the number and location of lymph nodes involved and the extent of metastatic spread.
- **cM**
  - Type: Value List
  - Permissible Values: ‘MX’, ‘M0’, ‘M1’
  - Description: CDE about the clinical M stage of the breast cancer situation (according to UICC).
  - Notes: It is important to note that different versions of the UICC staging system exist, and the criteria for each cM stage may vary slightly between versions. Accurate assessment of the cM stage requires comprehensive evaluation, including clinical examination, imaging studies (such as CT, MRI, PET scans), and sometimes biopsy or other pathological confirmation.
- **pM**
  - Type: Value List
  - Permissible Values: ‘MX’, ‘M0’, ‘M1’
  - Description: CDE about the pathological M stage of the breast cancer situation.
  - Notes: It is important to note that different versions of the UICC staging system exist, and the criteria for each cM stage may vary slightly between versions. Pathological confirmation of distant metastasis (pM) typically involves biopsy or other pathological evaluation of metastatic sites to verify the presence of cancer cells.

**Class ‘Criteria regarding location of tumor lesion’:**

- **Laterality of tumor lesion**
  - Type: Value List
  - Permissible Values: ‘left’, ‘right’, ‘both’
  - Description: CDE about the laterality of a tumor lesion
  - Notes: It is important to consider the context of the tumor's laterality in conjunction with other clinical, pathological, and genetic information to provide a comprehensive understanding of the disease and to guide optimal treatment strategies.
- **Tumor location**
  - Type: Value List
  - Permissible Values: ‘lateral’, ‘medial’, ‘central’, ‘lateral and central’, ‘medial and central’, ‘lateral, central and medial’
  - Description: CDE about the location of a tumor lesion within one breast
  - Notes: Assessing the exact location of a tumor can sometimes be ambiguous due to variations in breast anatomy, tumor size, and the presence of multiple tumor foci. Imaging techniques (such as mammography, ultrasound, and MRI) play a crucial role in determining tumor location but can yield varying interpretations. Additionally, anatomical landmarks used to define these regions may differ slightly between practitioners, leading to potential inconsistencies in classification.

**Class ‘General data about tumor lesion’:**

- **PrimaryOrRecurrect**
  - Type: Value List
  - Permissible Values: ‘primary’, ‘recurrent’
  - Description: Binary value about whether or not it is a primary or recurrent tumor lesion
  - Notes: The classification of a tumor lesion as 'primary' or 'recurrent' is critical for determining the treatment approach and prognosis. A 'primary' tumor refers to the initial occurrence of the cancer, while a 'recurrent' tumor indicates that the cancer has returned after treatment. Distinguishing between primary and recurrent tumors can sometimes be challenging due to ambiguities in clinical and pathological findings. For instance, differentiating between a new primary tumor and a local recurrence can be complex, especially in patients with a history of breast cancer.
- **associated DCIS of other tumor lesion**
  - Type: Value List
  - Permissible Values: ‘yes’, ‘no’
  - Description: Binary value about whether or not the tumor lesion is a DCIS and associated to another tumor lesion
  - Notes: The binary classification of ‘yes’ or ‘no’ indicates whether the DCIS is associated with another tumor lesion. Identifying and confirming the association between DCIS and another tumor lesion can be challenging. This assessment relies on thorough pathological examination of the breast tissue, including careful analysis of the margins and the spatial relationship between DCIS and invasive components. Variability in pathological techniques, interpretation criteria, and sampling methods can lead to inconsistencies in detecting and reporting associated DCIS.
- **Minimal resection margin**
  - Type: Number
  - Unit: mm
  - Description: Data value about the minimal resection margin of a tumor lesion
  - Notes: It measures the smallest distance between the edge of the tumor and the edge of the surgically removed tissue. There can be ambiguities and uncertainties in assessing the minimal resection margin. Factors such as tissue shrinkage during processing, variations in slicing and examining the tissue, and differences in pathological interpretation can all influence the measurement. Additionally, the definition of an adequate margin can vary depending on the type and location of the tumor, as well as institutional guidelines and clinical judgment.

**Class ‘tumor size’:**

- **Diameter (for ‘Diameter1’, ‘Diameter2’ and ‘Diameter3’)**
  - Type: Number
  - Unit: mm
  - Description: Data value about diameter of a tumor lesion.
  - Notes: There can be ambiguities and uncertainties in assessing the tumor diameter. Factors such as the method of measurement (e.g., imaging vs. pathological examination), the plane of measurement, and the potential for tumor distortion during specimen processing can all affect the accuracy of the diameter measurement. Additionally, intra-tumoral heterogeneity and irregular tumor shapes can complicate the assessment.
- **Modality of assessment**
  - Type: Value List
  - Permissible Values: ‘clinical’, ‘sonography’, ‘MRI’, ‘CT’, ‘PET-CT’, ‘histological’, ‘other
  - Description: Data value about the modality of assessment with which the diameter of a tumor lesion was determined.
  - Notes: It should be noted that the accuracy and reliability of tumor diameter measurements can vary significantly depending on the modality used.
